# HOW DO WE INDUCE LABOURS IN SPAIN? RESULTS OF THE INDUCESPAÑA SURVEY

**DOI:** 10.1101/2022.12.23.22283250

**Authors:** Pablo del Barrio Fernández, María del Carmen Medina Mallen, Rubén Alonso Saiz, Edurne Álvarez Suberviola, Cristina Álvarez Colomo, Emma Batllori Badia, María Begoña Dueñas Carazo, Elena Benítez Cano, Rut Bernardo Vega, Patricia Boza Novo, Laura Castro Portillo, Montserrat Comas Rovira, Lucía Díaz Meca, Camino Fernández Fernández, Irene Fernández Buhigas, Sara Fernández Menéndez, M. Belén Garrido Luque, Irene Gastaca Abásolo, Ruth Llano Fontela, Ivanna Llordella Sarmiento, Marina Llull Gomila, Rocío López Pérez, Marta Ruth Meca Casbas, Begoña Muñoz Abellana, Víctor Muñoz Carmona, Carmen María Orizales Lago, Rodrigo Orozco Fernández, María Caridad Ortiz Herrera, María Soledad Peña Salas, Sara Pérez Álvarez, Pilar Prats Rodríguez, Alberto Puertas Prieto, Linda Grace Puerto Tamayo, Silvia Rodríguez López, Susana Rodríguez Pradera, Cristina Sabench Carreras, Susana Soldevilla Pérez, María Suarez Arana, Elena Pintado Paredes, Mª Amparo Carrasco Catena, Óscar Martínez-Pérez

## Abstract

**Background:** Induction of labour (IDP) is the artificial initiation of labour with the goal of achieving a vaginal delivery. IDP is one of the most frequently performed obstetric procedures in the world. Recent data indicate a highly variable percentage of induction depending on the country.

**Methods:** A descriptive study was carried out on different aspects that are part of the ITP process through a survey prepared according to the Delphi method distributed among the 37 participating hospitals from June to October 2021.

**Results:** The mean induction rate was 30.6%. The average rate of caesarean sections was 21.4%. 75% of the centers had a rate higher than 20% and only in 1 center was it lower than 15%. In 11 centers they were not available to use misoprostol and in 4 centers they did not have oxytocin as PDI. Mechanical methods were available in 23 hospitals. In 16 centers they had a double Cook balloon; in 5 they used a Foley catheter and in 2 hospitals they used both devices. In 4 hospitals they used mechanical methods simultaneously with prostaglandins. In all but 5 centers, continuous monitoring was performed in low-risk pregnancies, at least for the first hour. In these 5 centers, windows of 20 minutes were carried out from the start of the induction

## INTRODUCTION

Induction of labour (IDP) is the artificial initiation of labour with the aim of achieving a vaginal birth. The Spanish Society of Obstetrics and Gynaecology (SEGO) defines it as the initiation of labour by means of medical or mechanical procedures, before the spontaneous onset of labour, with the aim of achieving delivery of the foetoplacental unit (1). Other international scientific societies define it as:

- artificial stimulation of uterine contractions before the spontaneous onset of labour to achieve vaginal delivery (ACOG) (2).
- initiation of contractions in a pregnant woman who has not gone into labour to assist her in achieving a vaginal delivery within 24-48 hours (SOGC) (3).
- artificial onset of labour (RCOG) (4).

In general, it is universally accepted that induction of labour (ITP) is indicated when the outcomes for the foetus, the mother or both are considered better than expectant management, i.e. waiting for spontaneous onset of labour (2).

Cervical ripening is fundamental for the evolution of labour and is a key factor in reducing the induction failure rate (IF). For the SEGO, cervical ripening is considered part of the IDP process and is defined as the set of biochemical and functional changes that occur in the connective tissue of the cervix, beginning in the first trimester of gestation and progressing to term, with the end result being changes in the cervix such as softening, shortening and dilatation (1).

Therefore, it can be said that cervical ripening is part of the IDP process and is indicated in cases where the conditions of the cervix are unfavourable at the beginning of the procedure.

Induction of labour (IPL) is one of the most frequently performed obstetric procedures in the world (1). The global frequency of IP has more than doubled between 1990 and 2012, rising from 9.5% to 23.3% (5).

Recent data indicate an induction rate of up to 35.5% in countries such as Sri Lanka (6) and 24.5% in the United States (7).

In Europe, the average rate of PID is 20%, but there are significant differences between countries. Thus, frequencies vary from 6.8% in Lithuania to 33% in Belgium according to the 2013 EURO-PERISTAT project report (8).

The same EURO-PERISTAT report of 2013 provides information from our country on the Valencian Community, with one of the highest rates at European level, 31.7% (8). We have not found reliable national figures, although very different rates are published for different national hospitals. In 2018, of the 250,704 deliveries attended in Spain, labour was induced in 83,624. The percentage of induced deliveries in public hospitals-SNS was 34.2%, continuing the upward trend of recent years (9).

Despite the extreme diffusion of the procedure, there are still many unanswered questions, or issues that have not obtained a unanimous consensus in the scientific literature.

### Medical and obstetric indications for termination of pregnancy by IDP

The only options for termination of pregnancy are induction of labour or caesarean section. Given the increased maternal risks associated with caesarean section, PID is the preferred option in the absence of contraindications to vaginal delivery. Current consensus indications include: post-term pregnancy, preterm and term premature rupture of membranes, hypertensive states of pregnancy, maternal DM, fetal growth restriction, twin pregnancy, chorioamnionitis, NIPPD, intrauterine fetal death.

### Contraindications for PID

There are a number of circumstances where the maternal and/or foetal risks associated with vaginal delivery, and therefore with induction, are greater than the risks associated with performing a caesarean section, therefore induction of labour is usually contraindicated (1): previous classical or corporal caesarean section, pregnancy after uterine rupture, pregnancy after transmural uterine incision with entry into uterine cavity, active herpes infection, placenta previa or vasa previa, umbilical cord prolapse or persistent cord procidence, transverse fetal status and invasive cervical cancer.

### Procedures available for induction of labour

A wide variety of methods are currently available to carry out PID. These can be divided into pharmacological and mechanical methods (10).

The most commonly prescribed drugs for labour induction worldwide are oxytocin and the synthetic prostaglandins (PGs) E2 (dinoprostone) and E1 (misoprostol). Oxytocin is the active substance we know most about because of its use over the years. It has been used alone, in combination with amniotomy or after cervical ripening with pharmacological and non-pharmacological methods.

In 2008 the Spanish Agency for Medicines and Health Products authorised the use of misoprostol 25 μg (Misofar^®^ 25 μg, Pfizer S.L. Madrid, Spain) in vaginal tablets for induction of labour and vaginal misoprostol 200 μg (Misofar^®^ 200 μg, Pfizer S.L. Madrid, Spain) for cervical ripening in gynaecological interventions. The bioavailability of PgE1 is three times higher vaginally than orally, and its peak action occurs around 45 minutes after administration (11).

The preparation of dinoprostone (PgE2) available in Spain is in the form of a vaginal pessary (Propess ^®^ Ferring Pharmaceuticals, Germany): it is a controlled-release vaginal device containing 10 milligrams of dinoprostone. The device releases 0.3 milligrams/hour for 24 hours. After 24 hours the device should be removed. Once removed, wait 30 minutes to initiate oxytocin (12).

Within the mechanical methods, there are single (Foley catheter) or double (Cook^®^ double balloon) intracervical balloons or probes. Theoretically, they work by direct physical pressure on the internal cervical orifice and produce the release of PG from the decidua, membranes and/or cervix. In addition, they promote biochemical and biophysical changes leading to cervical ripening and increased myometrial contractility (13).

#### BISHOP INDEX

In 1964, Edward Bishop established criteria for elective induction of labour that included parity, gestational age, fetal presentation, obstetric history and patient consent, as well as a scoring system for the cervix to help predict successful induction of labour. This pelvic scoring system, widely known as Bishop’s index, remains an important determination in predicting successful induction of labour. The score can be determined in a patient at the time of induction by digital cervical examination to determine whether cervical ripening is necessary prior to induction.

This index has a minimum of zero points and a maximum of 13 points. The scoring system uses cervical dilatation, position, effacement, cervical consistency and fetal station. A Bishop’s score of 8 or more is considered favourable for induction, or the likelihood of vaginal delivery with induction is similar to spontaneous labour. A score of 6 or less is considered unfavourable if induction is indicated and cervical ripening agents may be used in these cases (14).

The ideal procedure for PID is difficult to establish. This decision will depend on the clinical characteristics of the pregnant woman and foetus, the reason for induction and cervical maturity.

## OBJECTIVE

The main objective is to highlight the great variability in PID-related processes among a representative group of Spanish hospitals.

As secondary objectives we aimed to assess knowledge gaps in relation to induction procedures.

## SUBJECTS AND METHODS

This is a descriptive study on different aspects that are part of the ITP process by means of a Delphi survey that was distributed among the PIs of 51 national hospitals between June - October 2021.

### Survey design

Descriptive study conducted in Spain by a panel of experts following the online modified Delphi methodology with three rounds of participation. The project was coordinated by a scientific board of five experts led by PBF who designed and developed the Delphi consensus. The first round was held from 1 June to 21 June 2021, the second round from 21 June to 30 June 2021 and the third round from 1 July to 30 October 2021.

### Questionnaire

Initially, a PubMed and manual search was performed using specific keywords on four different topics: induction of labour, chronologically prolonged gestation, premature rupture of membranes, prostaglandins, twin pregnancy.

We designed the survey using the Delphi method with 3 evaluation phases with 3 concentric groups of experts. The questions were weighted until there was no significant variation in the experts’ opinions on the questions in the successive rounds. The steps we followed to reach consensus on the survey were as follows:

- Definition of the topic (induction of labour).
- A first panel of experts is selected.
- The first questionnaire was developed.
- We distribute the questionnaire and start the first round. Once the answers are obtained, the most relevant questions are established and the results are compared between centres.
- We defined the second panel of experts (researchers from a collaborative Emergency Obstetric Group whose work is primarily in the delivery room).
- Second round: we distributed and obtained the responses among another group of experts and developed and applied a new, more specific questionnaire based on the responses from the first round after expert consensus.

### Determining the degree of consensus in the first and second rounds

A 5-point Likert scale was used for responses to each item: strongly disagree, disagree, neither agree nor disagree, agree and strongly agree. After the first round, the percentage of each response for each item was determined. A second round was conducted to obtain consensus on those items where there were discrepancies.

A consensus of agreement was established when more than 80% of the participants responded with “agree” or “strongly agree” for the inclusion of the corresponding item. Similarly, a disagreement was defined when more than 80% of the participants responded “disagree” or “strongly disagree” to the corresponding item. When the two possible consensus options were not met, it was established that there was no consensus on the corresponding item.

We conducted a third round to reach a consensus with the second group of experts on the borderline discrepancies that remained after the second round and produced the final questionnaire.

After receiving the corresponding approvals from the Ethics and Research Committees, 37 of the 51 hospitals of the Spanish Group of Obstetric Emergencies that expressed interest in participating responded to the survey.

The remaining 14 centres did not receive Committee approval and did not participate in the study. The Spanish Obstetric Emergencies Group was formed in March 2020 to study the influence of COVID 19 and pregnancy with a grant from the Instituto Carlos III. This group registered cases of COVID 19 and pregnancy until April 2021 and has produced numerous publications. Heir to this group of 76 centres, the Spanish Obstetric Safety Group www.gesobstetrica.com was formed on 3 March 2022. It is in this group that this study, called InducEspaña, was initiated.

## RESULTS

### Hospital characteristics

The number of deliveries attended in the 37 participating hospitals in 2020 was 74876. Not all hospitals responded to all the questions asked. The mean induction rate was 30.6%; 65.7% of the centres (25/37) had induction rates above 30%. The mean caesarean section rate was 21.4%. Seventy-five per cent of the centres had a rate higher than 20% and only 1 centre had a rate lower than 15% (table 1).

**Table 1:**
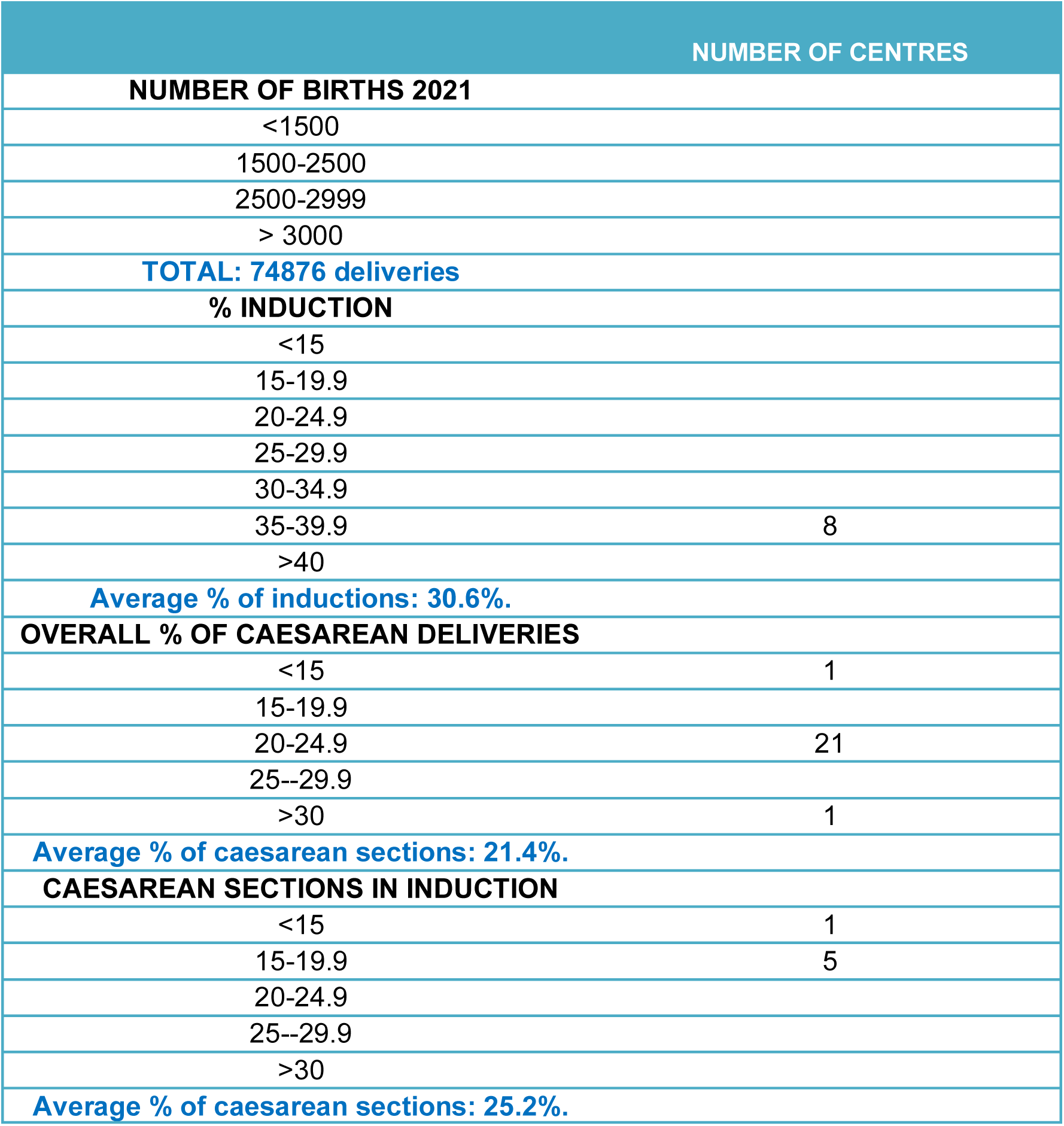
Characteristics of participating hospitals.

Regarding the indications for caesarean section and its relation to induction, the most frequent indication for caesarean section was dystocia (32.6%) followed by loss of foetal wellbeing (27.6%). The indication for caesarean section due to induction failure was 17.5% although in 3 hospitals (8%) the rate of induction failure was higher than 30%. The mean caesarean section rate for induced deliveries was 25.2%, although again there was great inter-hospital variability: in 1 centre this rate was less than 15%, while in 2 centres it was more than 35%. Sixty percent of the centres that responded to this section (18/30) had induction failure rates of less than 15%.

Only 3% of hospitals (1 centre) had overall caesarean section rates above 30%, compared to 26% (9 centres) with caesarean section rates above 30% in induced deliveries.

Overall, 73% of maternity hospitals had higher caesarean section rates for induced versus spontaneous onset deliveries, except in 2 hospitals (5%) where the caesarean section rate for induction was lower (figure 1).

**Figure 1:**
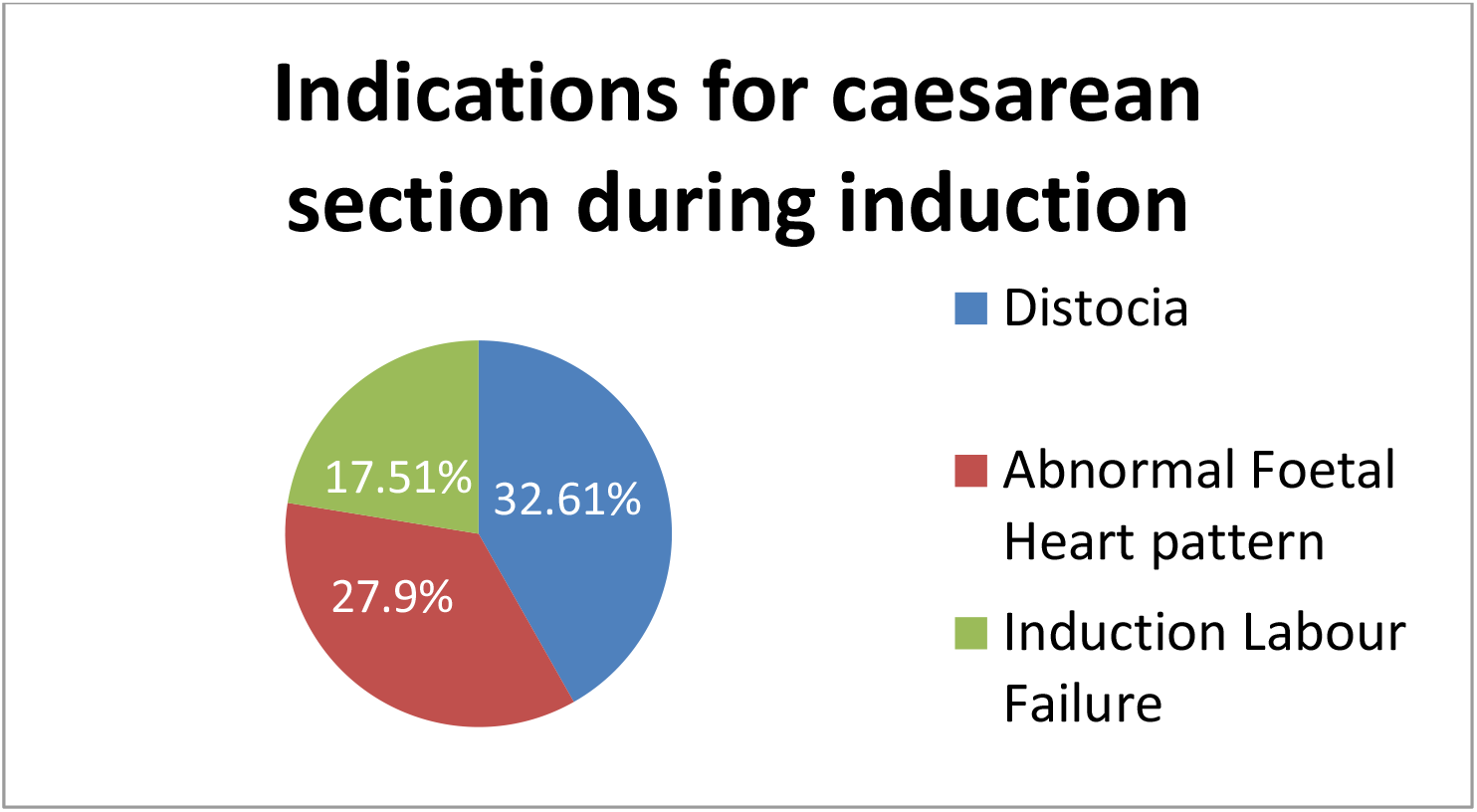
Indications for caesarean section during induction

The pharmacological methods available were prostaglandins (PgE1 and PgE2) and oxytocin. In 100% of the hospitals PgE2 was available for use. In 11/37 centres (29.7%) PgE1 was not available for use as an induction method and in 4 centres (10.8%) oxytocin was not used as an induction method.

In relation to mechanical methods, these devices were available in 23/37 of the delivery centres (62.1%). Specifically, in 16 centres (43.2%) the Cook^®^ balloon was available; in 5 hospitals (13.5%) the Foley catheter was used and in 2 hospitals (5.4%) both devices were used interchangeably. In 4 hospitals (10.8%) they use mechanical methods simultaneously with prostaglandins as a method of labour induction. The remaining 14 centres do not use these intracervical balloons.

In 44.4% of hospitals (12/27) the most commonly used induction method was dinoprostone; in 28.6% (6/21) misoprostol; in 13.6% (3/22) oxytocin, while in 6.6% (1/15) more than 50% of inductions were performed with mechanical methods (table 2).

**Table 2:** Induction methods used according to indication

| Induction of labor methods | Number of centres |
| --- | --- |
| Induction methods for all indications |  |
| Oxytocin | 33 |
| Prostaglandin E1 | 26 |
| Prostaglandin E2 |  |
| Mechanics | 23 |
| **Induction methods in RPM** |  |
| oxytocin |  |
| Prostaglandin E1 | 10 |
| Prostaglandin E2 | 18 |
| Mechanics | 0 |
| **Methods of induction in twin pregnancies** |  |
| Oxytocin |  |
| Prostaglandin E1 | 1 |
| Prostaglandin E2 |  |
| Mechanics | 5 |
| **Induction methods in previous caesarean section** |  |
| Oxytocin | 1 |
| Prostaglandin E1 | 0 |
| Prostaglandin E2 | 23 |
| Mechanics |  |
| **Induction methods in RIC/PEG** |  |
| Oxytocin | 0 |
| Prostaglandin E1 |  |
| Prostaglandin E2 | 23 |
| Mechanics |  |
| **Induction methods in GVP** |  |
| Oxytocin | 1 |
| Prostaglandin E1 |  |
| Prostaglandin E2 |  |
| Mechanics | 1 |

In all delivery centres the two most frequent indications for induction were prolonged gestation (PVG) and premature rupture of membranes (PROM). In 21/37 centres (56.7%) the first cause was PVG while in 16/37 hospitals (43.2%) it was PROM.

When the reason for induction was PVG, delivery was abdominal in 25.7% of cases, whereas when the indication was PROM, the mean caesarean section rate was 31.5%. The percentage of caesarean sections was higher in the PROM induction group (figure 2).

**Figure 2:**
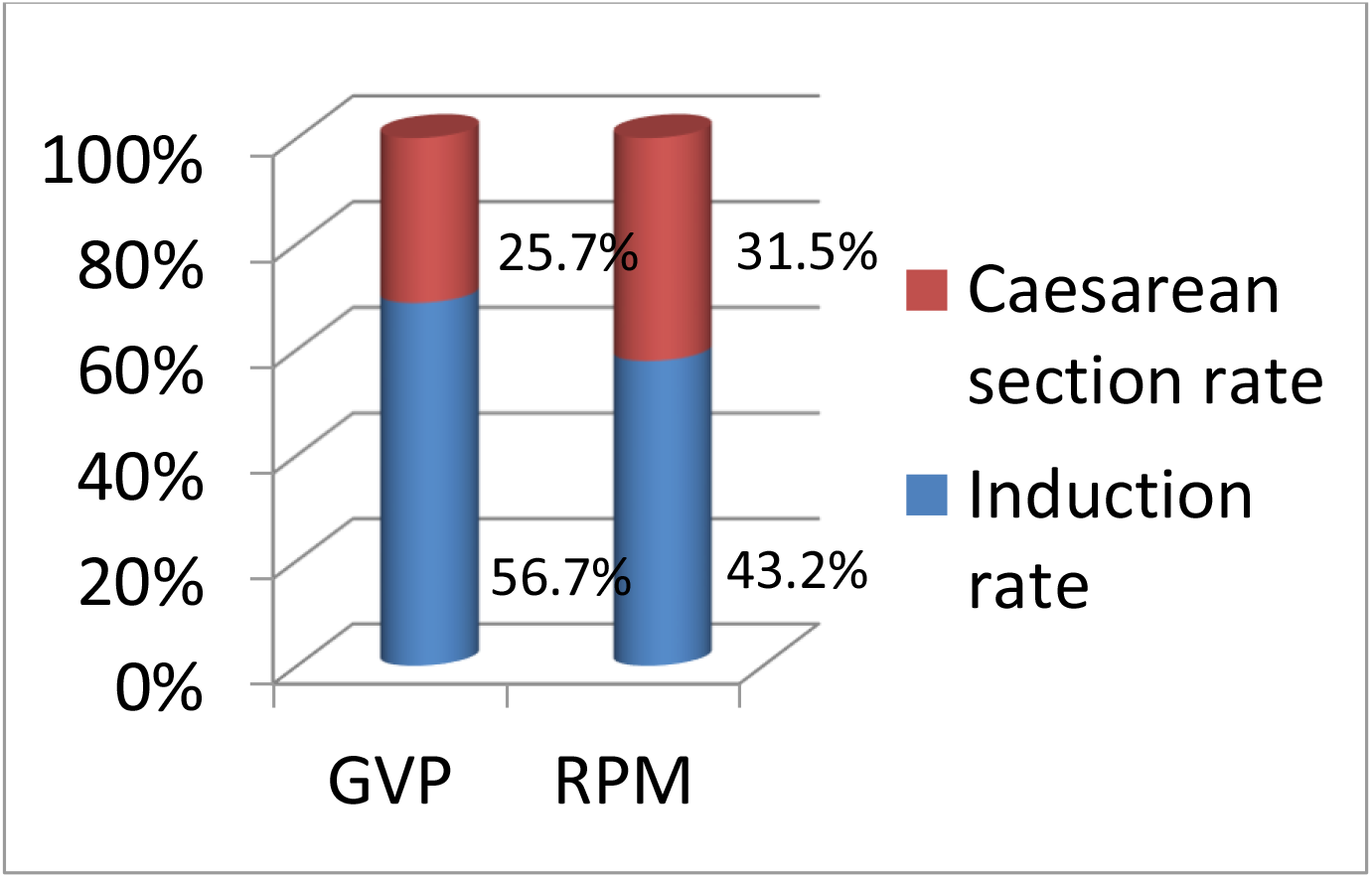
Most frequent indications for induction and percentage of caesarean sections.

When asked about the indication for induction in <u>premature rupture of membranes</u>, the hours of latency allowed before induction were up to 12h in 19 centres (51.3%); in 7 hospitals (18%) they waited up to 12-24h before induction and in 5 centres they delayed the start of induction up to 24-36 hours. In 4 centres (11%), they waited between 6-12 hours, which reflects an active induction behaviour in PROM. The most commonly used induction method in this situation was PgE2 (18 centres, 49%), followed by PgE1 (10 centres, 37) and finally oxytocin in 9 centres (24%).

The mode of gestational age at which prolonging pregnancies beyond <u>41 weeks</u> were induced was 41+3 weeks, a scenario that occurred in 10 of the 37 centres surveyed (27%). Two centres (5%) waited until 42 weeks to induce labour.

The most commonly used method to induce labour in hospitals with a prolonged gestation was PgE2 in 16 centres, followed by PgE1 in 13 centres, mechanical induction in only one centre and direct IV oxytocin in another centre. Specifically, only 31 of the 37 participating centres answered this question. The use of Bishop’s index to determine the method of induction in each case was always used in 17 of the centres, only a few professionals used it in 10 hospitals, none in 3 of the centres and 1 hospital used other methods to establish the method of induction. The cut-off point for this Bishop’s index to determine the induction method was <6 for 18 hospitals, <5 for 2 hospitals and in 8 centres there was no cut-off point, it was set at the discretion of the practitioner.

In 9 centres (24.3%) <u>macrosomia</u> was considered a criterion for induction; in 16 hospitals (43.2%) it was not a criterion for termination of pregnancy, while in 12 cases (32.4%) macrosomia was a criterion for induction only in diabetic pregnant women. In the specific case of <u>obesity</u>, in 89% of the centres (33/37) it was not a reason for induction.

In cases of <u>SGA/RCI foetuses</u>, dinoprostone was the most common method of induction in 23 cases (62%), followed by mechanical methods in 12 centres (33%) and in only 2 hospitals (5%) was misoprostol the method used.

The gestational age at which <u>twin pregnancies</u> were induced was 38 weeks in 19 hospitals (51.3%). Fourteen percent of the hospitals (5/37) induced between 38 and 39 weeks of gestation. As induction methods, dinoprostone was used first (28/37 centres), followed by mechanical methods (5/37 centres), oxytocin (3/37 centres) and finally misoprostol in only one hospital.

Hospitals requesting patients to sign an informed consent form for induction of labour in case of <u>previous caesarean section</u> were 25 (65.7%), while 11 (29.7%) did not require such a requirement. The gestational age at which labour was induced by previous caesarean section was the same as when there was no such history in 25 hospitals (65.7%) and in 11 of them (29.7%), the gestational age of induction of previous caesarean section was set at 41 weeks. The method of induction in these patients was mechanical in 13 of the centres (35%) and pharmacological in the remaining 24 centres (65%). Specifically, within the pharmacological methods, oxytocin was used in 1 centre and in 23 hospitals (62%) PgE2 was the elective induction method in patients with previous caesarean section (figure 3).

**Figure 3:**
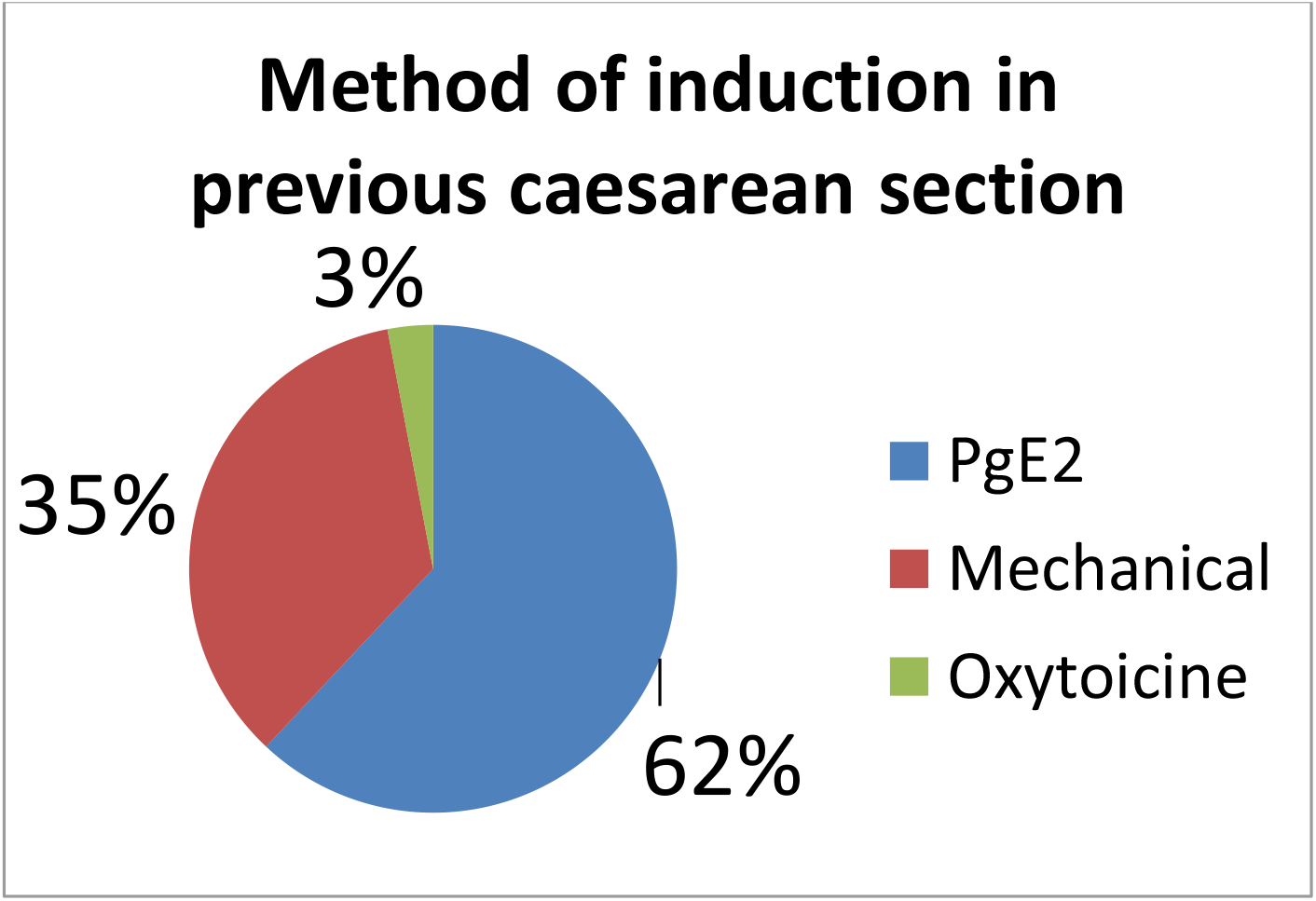
Induction method in previous caesarean section.

### Organisational aspects of induction of labour

In 33/37 centres (89.2%) the indications for induction are protocolised, the rest being discretionary. In 5 of the 37 hospitals no informed consent document was provided. In 27 centres, this document was drawn up at the centre itself; in 4 centres the document used was the one drawn up by the SEGO and in 1 hospital the hospital’s own document was provided together with that of the SEGO. In 27 cases, this document was given to the pregnant woman at the time the indication for induction was given, and in 7 cases the time of delivery was indifferent. In 3 centres, the document was given at the time of admission for induction. The preferred time of admission was first thing in the morning (32/37; 86.5%). Low-risk inductions were admitted on the ward in 22 hospitals (59.4%); in 12 cases they were admitted in the delivery room. In 1 centre, low-risk inductions were ambulatory and performed by mechanical methods. In all but 5 centres continuous monitoring was performed in low-risk pregnancies at least during the first hour. In these 5 centres, 20-minute windows were performed from the start of induction. In 29 hospitals analgesia was administered when requested by the pregnant woman irrespective of the stage of labour while in 8 (21.6%) analgesia was administered when requested by the pregnant woman, but only if she was in active labour (table 3).

**Table 3:** Organisational aspects of induction of labour

| ORGANISATIONAL ASPECTS | NUMBER CENTRES |
| --- | --- |
| Signed informed consent |  |
| Protocolised induction | 33 |
| Time of admission in the morning |  |
| Place of induction |  |
| - Inpatient ward |  |
| - Delivery room |  |
| - Outpatient | 1 |

*Table 3: Organisational aspects of induction of labour*
| Analgesia |  |
| --- | --- |
| - At maternal request, regardless of the stage of delivery | 29 |
| - At maternal request in active labour | 8 |

#### Prostaglandin induction

In 2 hospitals (5.4%) the <u>Dinoprostone pessary</u> was applied in a protocolised manner by the midwife; in 20 delivery rooms (54.0%) it was the obstetrician, while in 15 (40.5%) it was indifferent, it could be administered either by the obstetrician or the midwife in charge. The interval to the first examination, in the absence of contractions and with reassuring RCTG, was mostly at 12h (13/37), although in some centres it was done at 4-6h (11/37), in 8 centres (29.7%) at 8h and in 5 cases (13.5%) it was not explored unless the woman reported painful dynamics at some point in the process. The number of dinoprostone devices for induction was one in 33 hospitals; two in 2 centres and one every 24h in a row in 2 other delivery rooms. In case of hyperdynamia with foetal impact, 8 hospitals (21.6%) used O2 as part of the resolution measures. Specifically, 22 centres (59.4%) used ritodrine as a tocolytic, while 13 hospitals used atosiban (35%); the rest (5.6%) used one or the other interchangeably as resolution of hyperdynamia. When after 24h Bishop’s test was less than 7, 28 hospitals (76%) started amniotomy followed by oxytocin; 6 centres (16%) used oxytocin without amniotomy and only in 3 cases another induction device was placed again.

In relation to the <u>use of misoprostol</u>, the route of administration of this PgE1 was vaginal in 27 centres (73%) of cases. Only one hospital used the oral route in addition to the vaginal route. In 9 of the 37 hospitals surveyed, low-dose misoprostol was not available for induction of labour in their regular medical clinic. In 13 centres (35%) it was administered by the obstetrician; in the same percentage the professional administering it was indifferent and in 2 hospitals (5%) it was administered by the midwife. The maximum total dose of PGE1 was 125mcg in 13 centres (35%); in 6 hospitals (16%) 150 mcg and in 5 centres (14%) the maximum number of tablets was 4 (100mcg). The interval between doses was 4h in 19/27 hospitals; and in 7 of the centres 6 hours elapsed between one tablet and the next. Among the hospitals where misoprostol was available, 13 (46.4%) centres administered a maximum dose of 5 tablets (125 mcg). In case of hyperdynamia with foetal impact, the figures were similar to those for hyperdynamia with PgE2. 5 hospitals (18.5%) used oxygen in the resolution set; about 61% (13/29) used ritodrine as a tocolytic; and 31% (8/29) used Atosiban; the rest of the hospitals used Atosiban or ritodrine indifferently.

#### Mechanical methods

The volume of saline with which the balloon was filled was 80 ml in 68% of cases while in 7 centres it was 60 ml. Balloon inflation was progressive in 7 cases (30.4%) while in 15 centres (65.2%) it was progressive. In 1 case there is no fixed pattern of balloon or catheter filling. Regarding the use of analgesia, 18 centres (78%) did not use any method of pain relief during catheterisation. In 16 hospitals (70%) no traction was applied to the catheter. In 16 hospitals (69.5%) the low-risk induction was performed in a ward room; in 6 centres (26.1%) this occurred in a dilatation; and 1 of the hospitals had an outpatient protocol for low-risk balloon inductions. In 15 hospitals (65.2%) the catheter was kept in place for 12 hours while in 8 hospitals (34.7%) the time the catheter was kept in place was 24 hours. If after insertion of the cervical catheter and after 12-24 hours the Bishop’s test was less than 7, in 19 hospitals (82.6%) amniorrhexis and subsequent administration of oxytocin was performed and in 4 centres (17.4%) the next step was administration of prostaglandins. In only 2 centres (9%) was balloon tube insertion considered difficult. Despite this, 28 centres (75.6%) considered that specific training on the application of the balloon catheter was desirable. The same number of hospitals considered that this training would reduce inter-hospital variability and 6 centres (16%) did not answer this question.

## DISCUSSION

With the data obtained in our study we can see the great variability that exists between hospitals in our country when it comes to choosing the method of induction of labour.

The overall average caesarean section rate is similar to the average established in Europe according to the recently published EURO-PERISTAT report of November 2022. According to this review, there is a large variation in caesarean section rates, as well as differentiated trends. Caesarean section rates varied geographically in 2019, with lower rates in northern Europe (16.4 % in Norway, 16.6 in Iceland, 17.4 % in the Netherlands, 17.9 % in Finland) and higher rates in southern and central Europe (53.1 % in Cyprus, 44.4 % in Poland, 41.5 in Hungary, 34.8 % in Ireland). Spain, with 25.7 %, is very close to the European average (26.0 %). Between 2015 and 2019, twelve European countries have experienced a decrease in caesarean section rates, while in nine countries they have increased and in others they have remained stable. In Spain, the caesarean section rate has decreased slightly from 26.6 % in 2015 to 25.7 % in 2019 (15).

If we analyse the place where inductions are carried out, we can see that in almost all hospitals they are performed with hospitalisation and only one centre performs inductions with mechanical methods as an outpatient. American researchers have reported that preinduction with mechanical methods at home can be safe in appropriately selected patients, always being women with singleton low-risk gestation and excluding women with previous caesarean section, gestational hypertension or pre-eclampsia, pregestational diabetes, growth restriction or ruptured membranes (16-19).

In our setting the most common method of induction is dinoprostone, followed by misoprostol, oxytocin and mechanical methods respectively.

The most widely used system for establishing the induction method to date is still Bishop’s index. Multiple studies use it to divide patients into pre cervical ripening or direct IV oxytocin induction, with most of them setting the cut-off point at 6 (20).

A 2016 meta-analysis (21) comparing the use of misoprostol, dinoprostone and intracervical balloon for cervical ripening concluded that no method was clearly superior when taking into account both the 24h vaginal delivery failure rate, uterine tachysysystole with alterations in fetal heart rate (FHR) and caesarean section rate. Vaginal misoprostol and vaginal dinoprostone reduced the risk of failure of vaginal delivery within 24 hours, but increased the risk of uterine hyperstimulation with alterations in FHR. However, intracervical balloon was the least associated with uterine hyperstimulation with alterations in FHR. Finally, the use of oral misoprostol was associated with a lower risk of caesarean section. However, a meta-analysis recently published this November (22), involving 5460 women from 12 individual studies, has established that intracervical balloons, compared to vaginal prostaglandins, did not result in a significantly different rate of caesarean delivery (12 trials, 5414 women; crude incidence 27.0%; adjusted OR (aOR) 1.09, 95% CI 0.95-1.24; I2=0%), caesarean delivery for failure to progress (11 trials, 4601 women; aOR 1-20, 95% CI 0.91-1.58; I2=39%), or caesarean delivery for fetal distress (10 trials, 4441 women; ORa 0.86, 95% CI 0.71-1.04; I2=0%). Adverse perinatal outcome was lower in women allocated intracervical balloons than in those allocated vaginal prostaglandins (10 trials, 4452 infants, crude incidence 13.6%; aOR 0.80, 95% CI 0.70-0.0). 92; I2=0%). There was no significant difference in adverse maternal outcome (10 trials, 4326 women, crude incidence 22.7%; aOR 1.02, 95% CI 0.89-1.18; I2=0%).

According to Beckmann et al. if the cervix remains unfavourable after prostaglandin administration, the dose could be repeated or oxytocin could be started (23). Performing early amniorrhexis after the last dose of misoprostol or after withdrawal of dinoprostone shortens the induction time compared to waiting for spontaneous amniorrhexis (24,25). This is what happens in most hospitals in our study, where 76% of hospitals after 24h with a Bishop’s test <7 perform amniorrhexis and start intravenous oxytocin perfusion.

We can see that most hospitals that use misoprostol administer it vaginally and only one hospital also uses the oral route. This drug is rapidly absorbed both vaginally and orally (26) and there is no data to support one route as better than the other in terms of health outcomes. Looking at the results of our study for misoprostol, there is also variability in the dose used and the interval between doses. A meta-analysis conducted in 2021 supports the use of low-dose misoprostol for induction of labour and suggests that a starting dose of 25 mcg may be adequate, taking into account efficacy and safety (27).

If the decision is made to induce with dinoprostone, we can see that most hospitals use a single device per induction, relegating to a minority those hospitals that repeat with a new device if Bishop’s rate does not improve within 24h.

Of the hospitals surveyed, just over half (62%) have mechanical methods for induction of labour, the majority being the double Cook^®^ balloon. The fact that almost 40% of hospitals do not have this type of method is surprising, as studies to date associate mechanical methods with a lower risk of tachysysystole than prostaglandins (28,29). Moreover, according to the literature, only 6% of women who undergo cervical ballooning have an unfavourable cervix at 12 hours (28). In our setting, when this occurs, most hospitals (82.6%) proceed to perform amniorrhexis and subsequent oxytocin infusion. Early amniotomy (within the first hour after removal of the catheter) is associated with a higher success rate of vaginal delivery in the following 24 hours (31).

We have compared the overall caesarean section rate with the caesarean section rate in induced labour. The work of Grobman (32) emphasises that it is not an accurate comparison to assess the effect of induction of labour alone on the caesarean section rate; in his work comparing elective induction of labour in low-risk nulliparous women versus expectant management he concludes that the caesarean section rate is lower in the induction group. As is logical, the induction of labour group includes high-risk pregnancies in which the rate of operative delivery is naturally increased, which may lead to a bias when interpreting this type of results.

A decade ago, the administration of epidural analgesia was considered to be associated with an increased rate of caesarean section. A systematic review, published in May 2011 (33), coincides in its objective with this question. The authors in this review conclude that there is no increased risk of caesarean section or instrumental vaginal delivery if women start analgesia early (with cervical dilatation less than 3 cm) compared to starting analgesia at a later stage.

Intrauterine fetal resuscitation or intrauterine resuscitation is a set of non-operative techniques applied to the mother with the aim of improving fetal oxygenation by reversing the cause of the deteriorating fetal status, determined by a non-reassuring fetal heart rate (FHR) pattern. One such manoeuvre is the administration of oxygen to the mother when there is a repercussion of fetal heart rate tracing in prostaglandin-induced hyperdynamia. A recent pooled review (34) suggests that studies to date do not demonstrate an association between maternal oxygen and a clinically relevant improvement in UA pH or other neonatal outcomes, a practice that is routinely performed in 20% of our hospitals.

The success rate of vaginal (VP) delivery after caesarean section ranges from 72-76% (35-38), rising to 87-90% if there has been a previous vaginal delivery (38). For this to be possible, an appropriate balance between risks and chances of success is essential. Certain conditions make PV less likely, such as advanced maternal age, high BMI, high ultrasound-estimated fetal weight (EFW) and failure of progression of labour as the cause of the previous caesarean section (39). A retrospective cohort of women with a previous caesarean section as the only delivery (n=46,176) (40) describes the outcomes of induction of labour at 39-41 weeks and compares them with elective caesarean section and expectant management. They found a lower rate of caesarean section with induction, with no significant effect on perinatal mortality or uterine rupture, but with a higher risk of admission to the neonatal unit. The same study establishes that the risk of uterine rupture when there is only one previous caesarean section and the uterine incision was low transverse segmental is 0.31-0.47%. Although the absolute risk of uterine rupture after induction of labour is low, the relative risk, especially with the use of prostaglandins, is higher (35,36). Oxytocin, among pharmacological methods, represents the safest option for induction of labour (41). In a retrospective study published in France in 2017 (42), with a total of 269 patients, where prostaglandins were used as an induction method in patients with unfavourable cervix, it was concluded that dinoprostone was an effective procedure in patients with previous caesarean section, with morbidity comparable to other methods of labour induction. In our country, a study published in 2021 evaluating the efficacy and safety of dinoprostone in pregnant women undergoing PID with previous caesarean section found that this method did not appear to be associated with worse obstetric or neonatal outcomes compared to PID in pregnant women without previous caesarean section, but that it should be performed with special caution in this population group, using induction protocols and standardised doses of IV oxytocin due to the higher risk of uterine rupture (43).

These findings in the literature and in the routine clinical practice of 23 of our hospitals are extremely striking, since the same technical data sheet of the drug contraindicates its use if the patient has previously undergone an operation on the uterus, including delivery by previous caesarean section (12). Furthermore, the SEGO in its 2010 protocol for vaginal delivery after caesarean section, reflects a higher risk of uterine rupture, limiting its use with strict indications in immature cervices. From our results, we can say that the use of dinoprostone as an induction method is considered safe in patients with previous caesarean section.

Although the number of published studies is small, PgE1 (misoprostol) has been found to be associated with a very high risk of uterine rupture, with reported rates of up to 18% (44). Therefore, it should not be used in women with previous caesarean section (34,44) and more controlled studies are needed in this regard (46). Our published data are consistent with the literature and show that no hospital surveyed uses this method of induction in this group of patients.

Regarding indications for induction of labour and methods there are different positions in the literature. A systematic review (47) concludes that while for some indications induction of labour is clearly recommended, for some other common indications there is no strong evidence to support it. Overall, few RCTs have evaluated the various indications for induction. For conditions where there may still be a clinical trade-off regarding timing of birth, such as suspected macrosomia and high BMI, researchers and funding agencies should prioritise sufficiently powered studies that can provide quality evidence to guide care in these situations.

The development of a Clinical Practice Guideline by the scientific societies would contribute to the reduction of the variability found and improve the success rate of inductions.

## CONCLUSIONS

- There is great variability in indications, availability of methods, drug administration, resuscitative measures and timing of analgesia.
- There is variability in the use of mechanical methods of labour induction that are subject to an official technical sheet.

## Data Availability

All data produced in the present study are available upon reasonable request to the authors

## Notes

### Competing Interest Statement

The authors have declared no competing interest.

### Funding Statement

This study did not receive any funding

### Author Declarations

Ethical and Research Committee from Hospital Universitario de Getafe () 21/15.April 23rd 021 The 37 hospitals recognized this national certificated committee statement as lead center and were approved for the rest of the hospitals for the entire study

